# Identification of a specific APOE transcript and functional elements associated with Alzheimer’s disease

**DOI:** 10.1101/2023.10.30.23297431

**Authors:** Qiang Chen, Luis Aguirre, Huanhuan Zhao, Felix Borrego, Itziar de Rojas, Lingyan Su, Pan P. Li, Bao Zhang, Erzsebet Kokovay, James D Lechleiter, Harald H. Göring, Philip L. De Jager, Joel E. Kleinman, Thomas M. Hyde, Agustín Ruiz, Daniel R. Weinberger, Sudha Seshadri, Liang Ma

## Abstract

**INTRODUCTION:** The APOE gene is the strongest genetic risk factor for late-onset Alzheimer’s Disease (LOAD). However, the gene regulatory mechanisms at this locus have not been fully characterized.

**METHODS:** To identify novel AD-linked functional elements within the *APOE* locus, we integrated SNP variants with RNA-seq, DNA methylation, and ChIP-seq data from human postmortem brains.

**RESULTS:** We identified an AD-linked *APOE* transcript (jxn1.2.2) observed in the dorsolateral prefrontal cortex (DLPFC). The *APOE* jxn1.2.2 transcript is associated with brain neuropathological features in DLPFC. We prioritized an independent functional SNP, rs157580, significantly associated with jxn1.2.2 transcript abundance and DNA methylation levels. rs157580 is located within active chromatin regions and predicted to affect brain-related transcriptional factors binding affinity. rs157580 shared the effects on the jxn1.2.2 transcript between European and African ethnic groups.

**DISCUSSION:** The novel *APOE* functional elements provide potential therapeutic targets with mechanistic insight into the disease’s etiology.

## INTRODUCTION

Alzheimer’s disease (AD) is a devastating neurodegenerative disease characterized pathologically by the accumulation of amyloid-β plaques and tau tangles, which leads to neuronal cell death and cognitive impairment. Most AD cases are non-Mendelian and late-onset (> 65 years old), and there is only limited treatment to slow down cognitive decline (e.g., lecanemab^1^), making AD the leading cause of mortality in the aging population^2^. African Americans remain underrepresented in AD research, despite the prevalence of AD being possibly double in frequency in AA compared to European Ancestry individuals^3^.

The human *APOE* protein has three common isoforms defined by two single nucleotide polymorphisms (SNPs) that reside in the coding region of exon 4. Of note, the apolipoprotein E gene (APOE) epsilon 2 (APOE2) and epsilon 4 (APOE4) alleles are two major genetic risk factors for late-onset AD. Compared to the commonest genotype (homozygous genotype comprising two copies of the APOE epsilon 3 APOE3/3), people carrying two APOE4 alleles (homozygotes) are at the highest risk^4^. Yet, there is no therapeutic intervention to reduce this risk of APOE4 carriers. Therefore, uncovering and understanding biological effects regulating the expression of APOE isoforms might contribute to the control of this important AD risk factor.

Recently, we performed genome-wide association studies (GWAS)^5^ and identified many AD-risk SNPs within the *APOE* gene region (**Supplementary Fig. S1**). However, most of these identified signals are in non-coding regions and are in complex linkage disequilibrium (LD) with other variants, including the SNPs encoding for the protein isoforms of APOE. Although we suspect the existence of additional variants modulating the risk of APOE isoforms, the complexities within the locus might present difficulties in elucidating their potential modulation of AD-related risk alleles. Cis-acting expression quantitative trait loci (eQTLs) studies might help to improve our understanding of the mechanisms of AD-associated variants in the regulation of the *APOE* gene expression^6,7^. Interestingly, a splicing variant of *APOE* mRNA with intron-3 retention, a long non-coding RNA, was found to govern *APOE* gene expression in neurons^8^. Furthermore, this non-coding RNA of *APOE* is more abundant in AD with more severe tau and amyloid pathological burden^9^. In contrast, we still do not know the roles of each *APOE* protein-coding transcript in AD pathogenesis. A study between *APOE* transcription and AD pathology has been attempted in AD brains from the superior temporal gyrus, but no significant correlation was determined^10^.

Another challenge is to understand the specific mechanism(s) by which variations at the *APOE* locus alter risk, including DNA methylation, chromatin activity, transcriptional factors (TFs) binding, and their interaction with SNPs and specific *APOE* transcripts. Level changes of DNA methylation were observed in AD subjects in the *APOE* CpG islands within exon 4 compared to age-matched controls in brain tissue^11^. Chip-seq of histone marks has been generated at the *APOE* locus from several studies^12^. However, how common risk alleles influence the epigenetic elements in AD remains largely unknown.

The present study aims to connect common AD risk alleles at the *APOE* locus with transcript(s), CpGs, and active chromatin regions by combining available human postmortem brain high-throughput functional genomics data. We leveraged two large human autopsy brain cohorts collected by the Religious Orders Study/Memory and Aging Project (ROSMAP)^13^ and the Lieber Institute for Brain Development (LIBD)^14^. Overall, we deepen our understanding of genetic and epigenetic regulation of *APOE* in the postmortem brain and provide a foundation for formulating mechanistic hypotheses for the variants within APOE associated with AD risk.

## RESULTS

To elucidate the mechanism of AD risk variants and its connections with transcriptomic, genetic, and epigenetic features within the context of AD, we harnessed the power of available multi-omics datasets sourced from diverse brain regions and two ancestries (**Fig. 1A**). It is noteworthy that while certain facets of this dataset have previously been analyzed in studies exploring brain phenotypes^15,16^, these earlier investigations predominantly emphasized genome-wide patterns. In contrast, our current study is distinct in its focus to unravel the intricate regulatory mechanisms operating within the *APOE* locus. As a novel contribution, we present, for the first time, compelling associations between AD-associated risk SNPs and important functional elements at the *APOE* locus.

**Figure 1.**
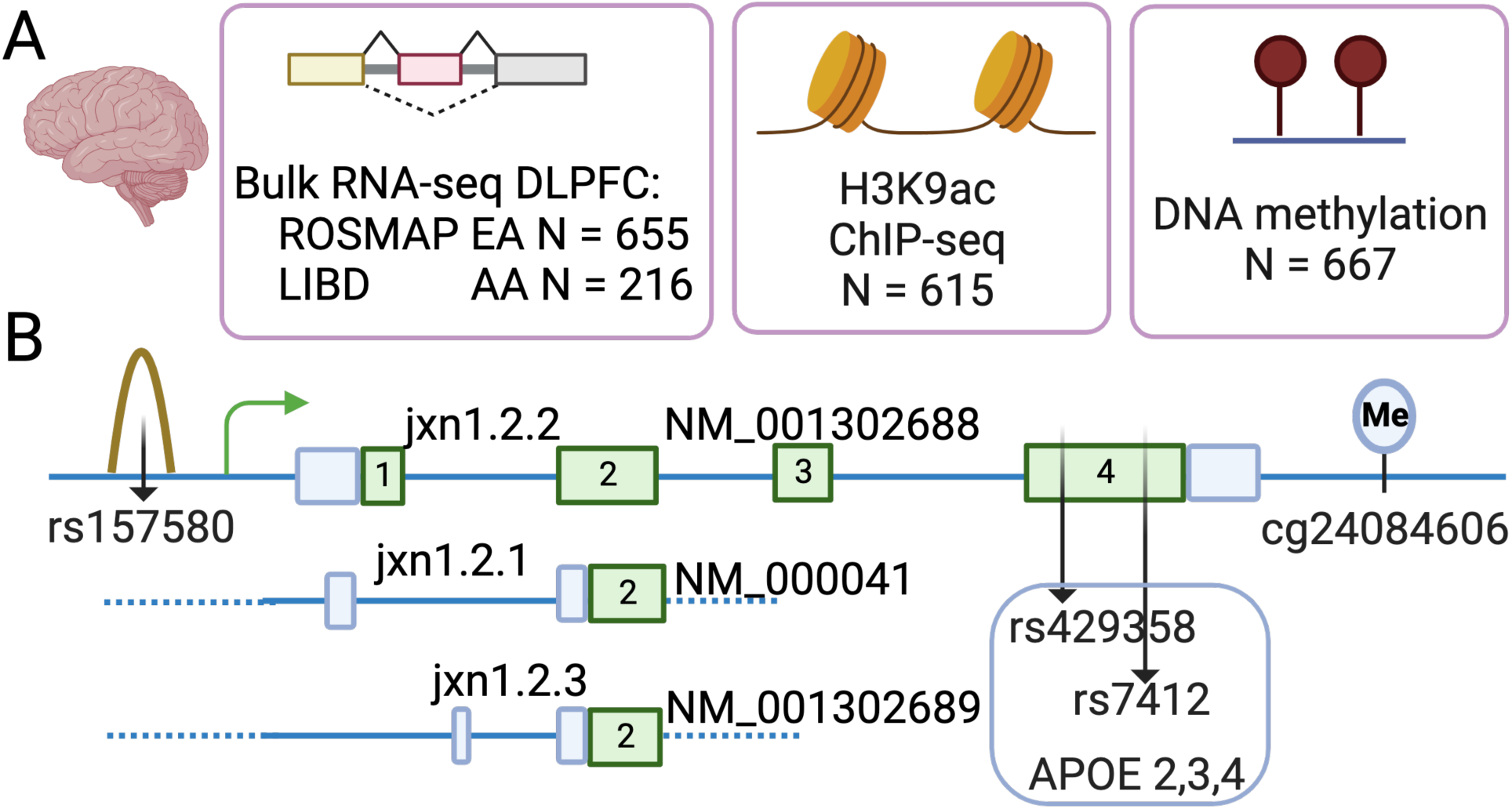
Overview of APOE study in human postmortem brain (A) and novel AD functional elements (genetic, transcriptomic, and epigenetic elements) and their relative position at the APOE locus (B). Brain collection: ROSMAP, The Religious Orders Study and the Memory and Aging Project; LIBD, Lieber Institute for Brain Development. Ancestry: EA, European Ancestry; AA, African American. Brain region: DLPFC, dorsolateral prefrontal cortex.

Our investigative journey commenced with a comprehensive exploration of the *APOE* locus, extracting transcriptomic, methylation, and histone modification features from the ROSMAP dorsolateral prefrontal cortex (DLPFC) dataset (see data availability). Serving as our cornerstone, this brain region formed the basis for probing *APOE* gene expression, encompassing bulk tissue RNA-seq (n = 655), histone modification through H3K9ac ChIP-seq (n = 615), and DNA methylation utilizing the 450K Illumina array (n = 667). Applying a congruent methodology, the LIBD dataset (see Methods) became another vital resource for investigation. With the DLPFC brain region at its core, this dataset facilitated the accumulation of additional bulk RNA-seq data from African Americans (n = 216).

Because the vast majority of genes are regulated within an enhancer’s chromosomal position (cis-regulation), we limited our transcriptional mechanism studies to 2 Mb^17^ spanning the *APOE* gene. To select potential functional variants in the selected region, we extracted the genotypes of 6,428 high-quality SNPs from ROSMAP whole-genome sequencing data, and 10,838 SNPs from LIBD African for downstream analysis.

### *APOE* jxn1.2.2 transcript is uniquely linked to specific AD risk-associated alleles in the *APOE* region

To pinpoint *APOE*’s mRNA transcripts within specific gene regions, we employed an expression feature known as exon-exon junctions. This approach effectively tags specific transcripts, enhancing our ability to quantify them with a heightened degree of precision and specificity, as demonstrated by our recent postmortem brain studies^18–20^. Following the reads alignment and quality controls, our efforts yielded three distinct splicing junctions connecting exon 1 and exon 2, alongside a common junction spanning exon 2 and exon 3, as well as another common junction bridging exon 3 to exon 4 (**Fig. 1B**). Consequently, our focus homed in on the junction linking alternative exons 1 and 2, a pivotal choice given its capacity to delineate diverse *APOE* transcripts. Then, we combined the *APOE* gene expression information with genomic variants previously selected with the aim to identify the SNPs associated with the levels of the *APOE* transcripts identified. Specifically, we examined the association of selected variants with the global abundance of *APOE* expression (combining reads of all transcripts identified) as well as the abundance of each different spliced isoform. To this end, we initially adjusted the dependent variables by ancestry and potential batch effects by regressing the expression levels with five ancestry principal components (PCs) derived from sequencing data and K PCs to correct potential batch effects detected by sva^21^ (detailed in Methods). The Adjusted expression levels were then fit to SNP genotypes, covarying for sex and diagnosis, using an additive linear model implemented in TensorQTL^22^. Across the RNA-seq datasets (**Fig. 1A**), we identified an average of 57k SNP-gene pairs and 5M SNP-junction pairs at the *APOE* locus, about 6k and 12k cis-eQTLs at gene and junction levels with a false discovery rate (FDR) < 0.05.

To link the *APOE* transcripts-associated variants (eQTLs) to AD risk alleles, we co-localized observed eQTLs with AD GWAS^5^ SNPs (**Supplementary Fig. S1**). The integration yields an average of 355 SNP-gene pairs and 586 SNP-junction pairs with genome-wide significance for AD risk (p < 5e-8) and FDR-significant for eQTL analysis (FDR < 0.05). Importantly, we uncover that a particular junction between alternative exon 1 and exon 2 (named jxn1.2.2 and tagging the *APOE* transcript NM_001302688) is the top hit junction at the *APOE* locus co-localizing with variants associated with AD-risk (p = 1.71e-13) (**Fig. 1B and 2A**). We didn’t observe statistical significance between AD risk variants (GWAS p < 5e-8) and other *APOE* transcripts (jxn1.2.1 and jxn1.2.3) or *APOE* gene-wide expression level (**Fig. 2B**).

**Figure 2.**
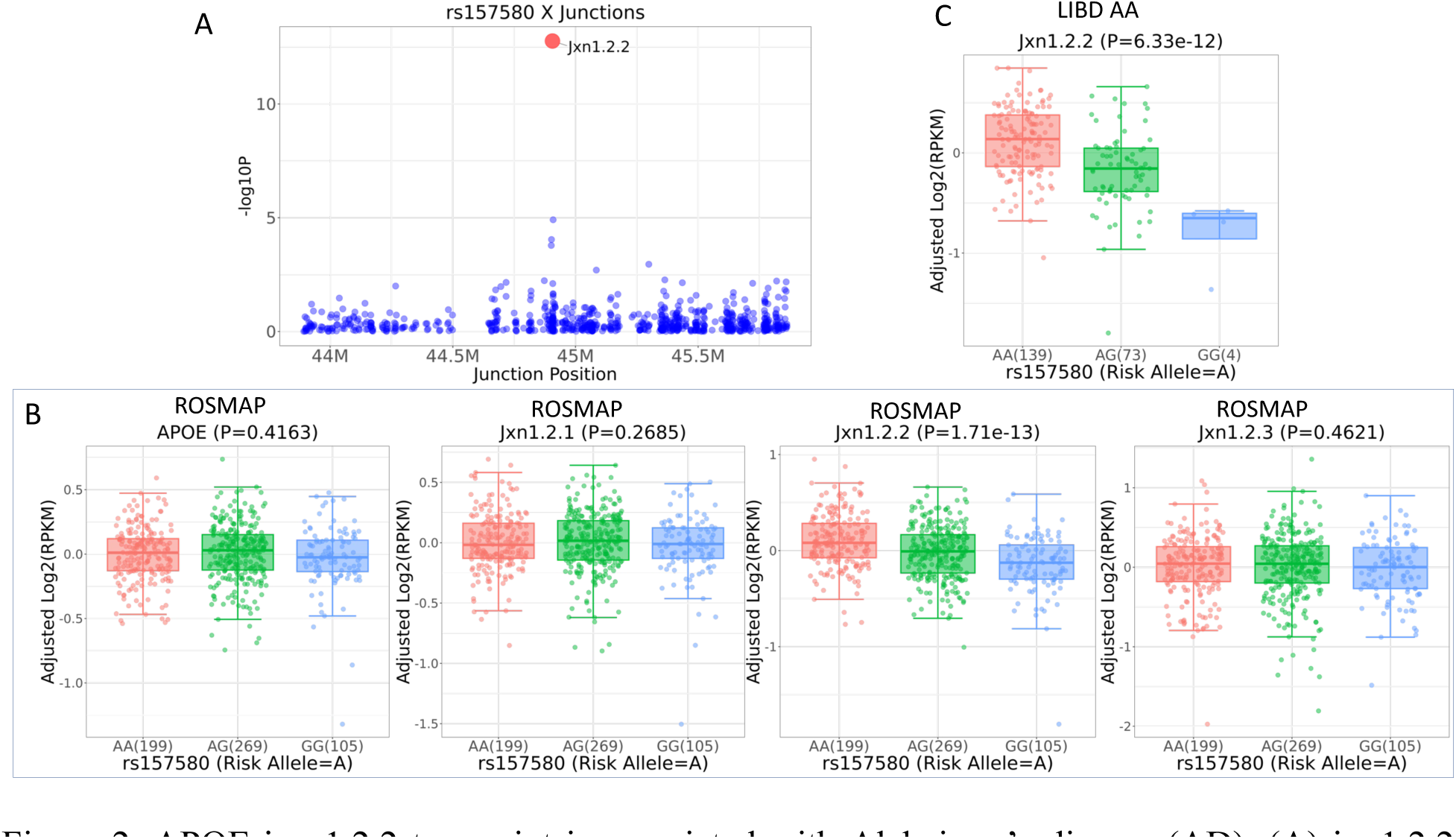
APOE jxn 1.2.2 transcript is associated with Alzheimer’s disease (AD). (A) jxn1.2.2 expression (red) is the top hit compared to other transcripts at the APOE locus (blue). (B) The association of AD risk SNP, rs157580, with APOE gene level and its 3 transcripts (jxn1.2.1, jxn1.2.2, and jxn1.2.3) in ROSMAP European ancestry. (C) Association of jxn1.2.2 and AD risk SNP in LIBD African American (AA).

To assess the potential influence of ancestry on the relationship between *APOE* jxn1.2.2 transcripts and AD genome-wide significant risk alleles (‘AD alleles’ hereafter), we also conducted an analysis of RNA-seq data from the LIBD African ancestry brain DLPFC collections, and this association persists (**Fig. 2C**), suggesting a significant link between *APOE* jxn1.2.2 transcripts and AD alleles in samples from two different ancestries. Because we analyze European and African ancestries separately, the local ancestry may not be representative for the whole heterogeneity among ancestries. Instead, we performed global ancestry analysis using the Identity by Decent (IBD) test and Principal Component Analysis (PCA) by integrating genotype data of ROSMAP and LIBD separately with HapMap3 populations (see Methods and **Supplementary Fig. S2**).

The gene structure of *APOE* consists of four exons, with the two SNPs (rs429358 and rs7412 located in exon 4) determining the three common protein isoforms of the *APOE* gene (**Fig. 1B**). To determine if the association of AD alleles with jxn1.2.2 transcript is independent of the APOE2,3,4 alleles, we performed the conditional analysis by including the APOE2,3,4 defining SNPs as co-variants, and found the results were not influenced in independent datasets: ROSMAP European and LIBD African populations (**Supplementary Fig. S3**). To further define the independent effects of our candidate AD alleles on APOE jxn1.2.2 expression from APOE genotypes, we performed epistasis (statistical interaction analysis), and we did not observe significant interactions between our candidate AD alleles and the APOE4 and APOE2 alleles (**Supplementary Table S1**), indicating the association between jxn1.2.2 expression and our candidate AD-risk alleles is not influenced by APOE4. The independent expression of jxn1.2.2 transcript was further supported by the lack of association between APOE2,3,4 determining SNPs and jxn1.2.2 expression (**Supplementary Table S1**).

### *APOE* jxn1.2.2 transcript expression levels are associated with AD pathology in DLPFC

To explore the role of *APOE* transcripts abundance in AD, we compared its expression level between AD and controls using different AD endophenotype Braak criterion to evaluate the density and distribution of neurofibrillary tangles (NFT)^23,24^. At the gene level by combining all transcripts, the *APOE* expression was not differentially expressed in DLPFC brain region. At the single transcripts level, by analyzing the three transcripts separately, we found that jxn1.2.2 transcript was differentially expressed between AD and controls compared to other *APOE* transcripts (jxn1.2.1 and jxn1.2.3) in DLPFC (**Fig. 3**).

**Figure 3.**
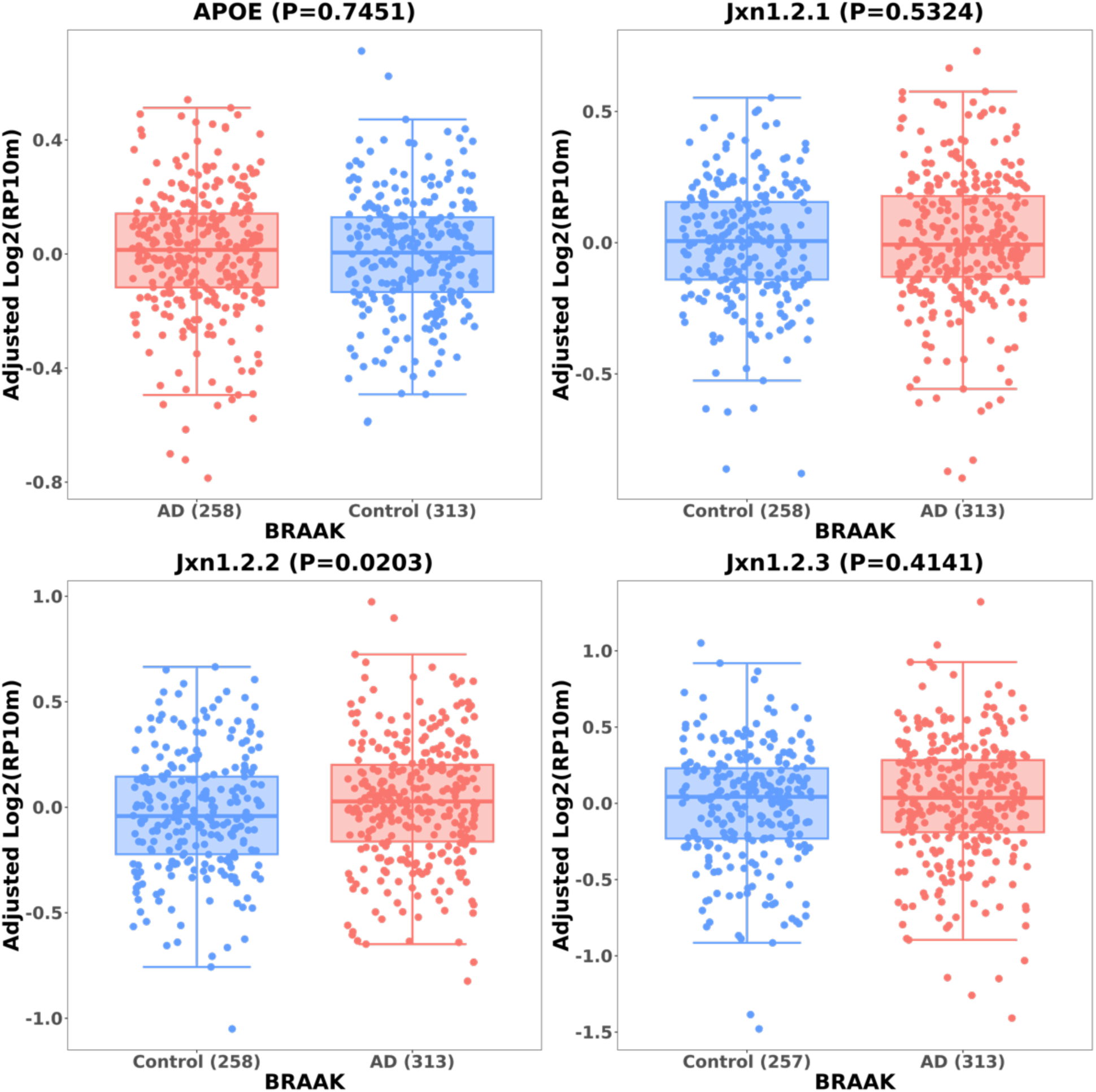
Differential expression of APOE at the gene level and transcripts level between AD and controls in BRAAK diagnosis.

### Identifying functional SNPs using epigenetic data from brain tissues

To identify potential regulatory SNPs in the *APOE* region, we carried out a rigorous statistical effort to identify CpGs spanning the *APOE* region. We obtained 777 CpG sites and performed association analysis between 7,335 SNPs and methylation levels in selected epigenetic features (mQTL). After filtering with mQTL FDR < 0.05, we obtained 5,029 SNPs and 312 CpG sites. Subsequently, to link the DNA methylation with AD, we integrated selected CpG sites with AD variants and eQTL results. We observed significant impacts of AD alleles of rs157580 on DNA methylation cg24084606 (p = 1.3e-9) (**Fig. 4A**). To determine whether the effect of DNA methylation can be modified by the APOE4 allele, we performed conditional analysis by including the APOE2,3,4 defining SNPs as co-variates, and found the results were not influenced (**Fig. 4B**). We also checked for statistical interaction between methylation levels and AD alleles were influenced. As expected, we did not observe significant interactions between our candidate AD alleles and APOE4 on the DNA methylation levels (**Fig. 4C**). Consistent with the independent relationship, we found that APOE2,3,4 determining SNPs are not associated with our prioritized CpG methylation levels.

**Figure 4.**
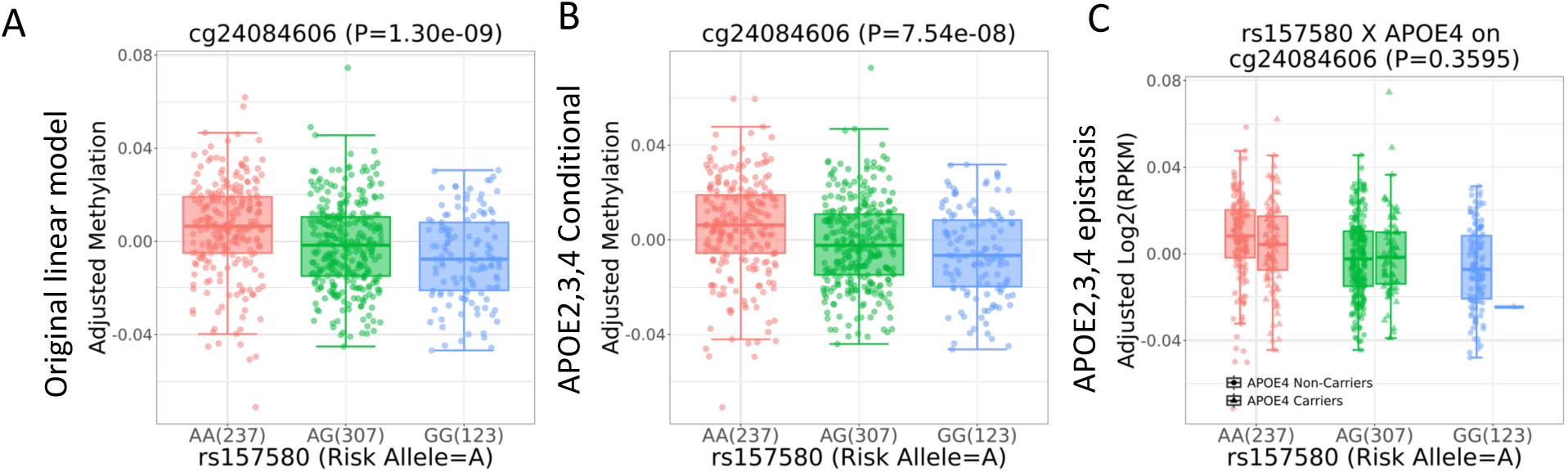
Genotypic impact of rs157580 on DNA methylation levels of cg24084606 in ROSMAP DLPFC brain tissue. (A) Association of the candidate AD risk SNPs with CpG sites. (B) The association of the AD-allele-linked CpGs is not affected by the APOE4 allele by Conditional analysis. (C) No interaction between APOE4 and rs157580 on DNA methylation levels.

ChIP-seq experiments can determine which chromatin regions are actively involved in gene transcription. Here we carried out several steps to prioritize SNPs within active chromatin at the *APOE* locus: First, we identified that rs157580 is located within active chromatin regions (**Fig. 5A**). Second, most enhancers exert their regulatory function through the binding of TFs. Thus, we performed an in-silico search of the DNA sequence of the SNP for putative TF binding sites using Motif Scan and Enrichment Analysis (MoSEA). Third, we reviewed the literature and found motifs affected by SNPs that were reported to be involved in neuronal function. rs157580 was predicted to be located within binding sites of EGR4 and vitamin D receptor (VDR) (**Fig. 5B**).

**Figure 5.**
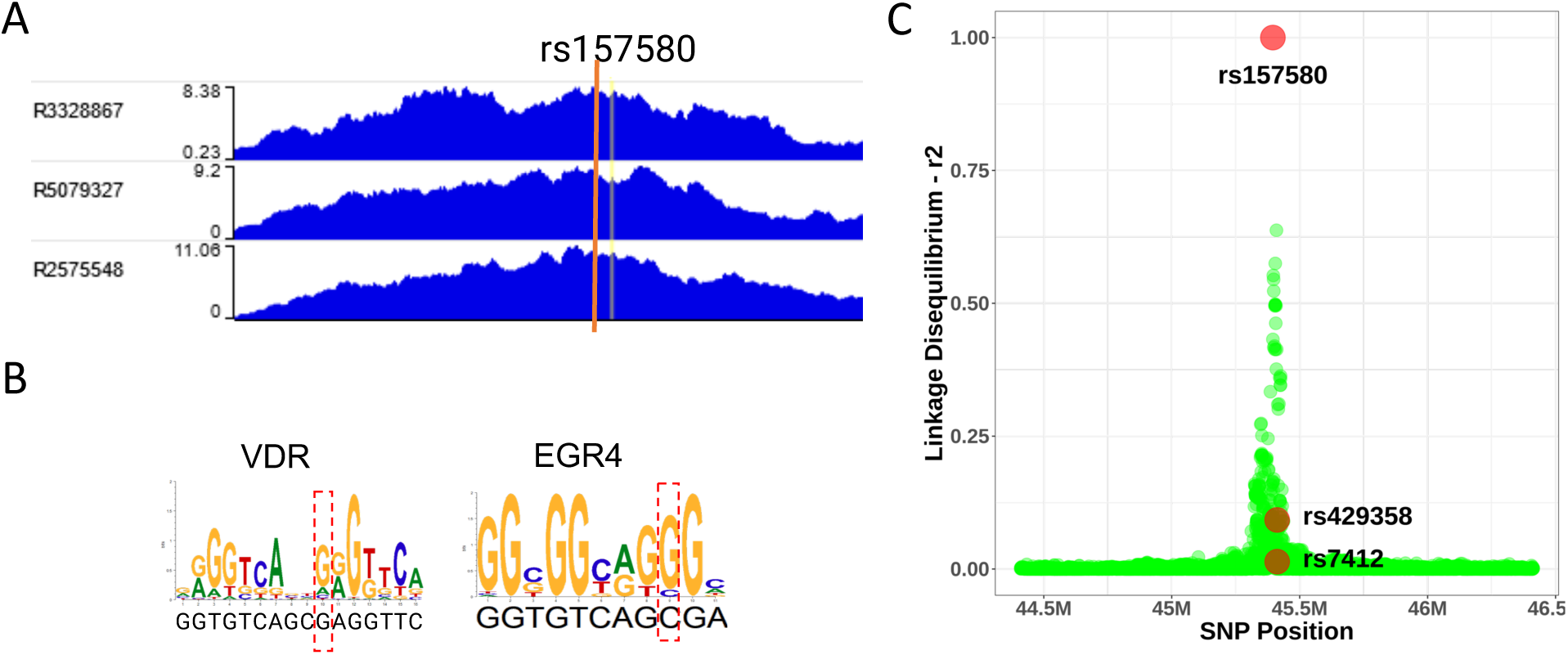
rs157580 is located within active chromatin and affects transcriptional factors (TFs)’ binding affinity. (A) rs157580 is co-localized with H3K9ac ChIP-seq peak from human postmortem brains. (B) Recognition sites of TFs involved in Alzheimer’s disease are influenced by rs157580. The red dash box indicates the binding site of each SNP. (C) Linkage disequilibrium of rs157580 with other SNPs spanning APOE, including the two APOE2,3,4-determining SNPs.

rs157580 is not significantly associated with global *APOE* levels in European and African populations across our datasets. However, they were associated with the jxn1.2.2 transcript (FDR < 0.05). While rs157580 is associated with jxn1.2.2 expression levels in European, it is also significantly associated with jxn1.2.2 expression levels in African, indicating the shared regulatory mechanisms for both ancestries. Importantly, rs157580 may represent partially independent meQTLs associated with AD risk, according to the weak linkage disequilibrium with the common AD-risk polymorphisms (rs7412 and rs429358 defining the APOE2,3,4 alleles, **Fig. 5C**). Furthermore, CSF Amyloid-beta 42 (Aβ42) and phosphorylated tau (pTau) are two major proteins implicated in the AD pathological process that can be assayed. We studied the genetic effects on CSF Aβ42 and pTau levels in a total of 13,116 individuals using GWAS data^25^. We found that rs157580 is associated with both biomarkers in CSF (p = 4.37e-74 and 1.97e-58 separately) (**Supplementary Fig. S4**).

## DISCUSSION

The APOE2,3,4 are not the only genetic risk factors for AD. Indeed, GWAS studies^5^ have identified numerous potential AD risk SNPs. However, the molecular mechanism of most AD loci remains largely elusive. Despite *APOE* has long been a widely investigated gene since the identification of its association with lipid levels and AD, the biological mechanisms behind these associations are unknown. Many studies have reported the relationship between APOE2,3,4 protein isoforms and AD-related traits, such as impairing synaptic repair and plasticity^26^; increasing beta-amyloid aggregation^27–29^; increasing formation of neurofibrillary tangles; and decreasing metabolic activity of neurons^30^. These phenotypes have been largely attributed to APOE2,3,4 protein isoform biochemical properties that differ by single amino acid substitutions constituted by alleles of rs7412 and rs429358^31^. Indeed, beyond the overt differential molecular bending of APOE2,3,4 isoforms and subsequent alterations in lipidation capacity^32,33^, there is limited evidence supporting functional variants at this locus modulating full-length APOE isoforms.

Here, we provide evidence of additional functional elements at the *APOE* locus that may contribute to the mechanism of action of the *APOE* locus in AD and related phenotypes. We leveraged data from multiple large population-based cohorts of human postmortem brains in diverse ethnic groups. Our study offers insights into the genomics elements controlling *APOE* expression in the brain, but the pathological relevance of observed *APOE* transcripts by including/excluding exons and their regulatory mechanism will need additional clarifications in the future. Similar to our work in *SNX19*^19^ and *CYP2D6*^18^ genes, we demonstrate that a careful analysis of postmortem brain data can identify specific *APOE* gene transcription mechanisms associated with AD-risk alleles. Our results prioritize specific domains between exon 1 and exon 2 in the protein that contain the functional domain that might influence AD risk. The data made us aware that the AD susceptibility signals can also be masked in gene expression analysis, and that the focus on individual transcripts is absolutely crucial to understanding APOE mechanisms operating not only in the brain but also in other tissues expressing this pleiotropic gene. Furthermore, pinpointing additional functional mechanisms modulating causal common variants at the *APOE* region and elucidating their roles in AD susceptibility might contribute to delineating therapeutic strategies for controlling this important susceptibility factor. Unfortunately, controlling APOE-associated risk remains a major challenge of dementia research. Our results therefore refine our understanding of the *APOE* locus and suggest that genetic variant affecting APOE regulatory motifs might have independent effects influencing AD susceptibility.

Strengths of this study include the use of the ROSMAP cohort in our main analyses, and extended in the LIBD cohort and its connection with large meta-GWAS of AD risk. The ROSMAP brain collections are unique in terms of their longitudinal nature, and in the ages of the subjects involved. This study is also strengthened by identifying the potential pathogenic role of *APOE* jxn1.2.2 transcript, and replicating it in the additional cohort with different ancestry. Importantly, this transcript is also a risk expression feature in the African ancestry population. The *APOE* jxn1.2.2 transcript was differentially expressed between AD and controls and in the DLPFC brain region. The DLPFC is a region affected by amyloid-β pathology relatively early as it spreads throughout the neocortex^34^. The accumulation of tau pathology progresses stereotypically captured by the braak stages^35^, and the DLPFC displays an accumulation of neurofibrillary tangles containing tau typically when individuals begin to be symptomatic. Thus, both pathological amyloid-β and tau accumulate in the DLPFC in AD, and we use quantitative measures of these pathologies to enhance our power in discovering the molecular features that are associated with these pathologies. However, we feel that characterizing more brain regions is very necessary to understand its potential role in AD pathogenesis and its connection with mature APOE protein isoforms.

A major finding of this study is that the *APOE* jxn1.2.2 transcript might differentially contribute to AD risk compared to other alternative transcripts. We found that AD alleles are specifically associated with enhanced jxn1.2.2 expression. Consistently, we also found that upregulation of *APOE* jxn1.2.2 transcript is associated with AD hallmark NTF. Those findings support our hypothesis that AD-linked *APOE* transcript signals can be masked in analysis at the gene level. Because this transcript is partially affected by the APOE2,3,4 alleles, it might be an important additional factor within this *APOE* region. To the best of our knowledge, this is the first study to pinpoint this AD-linked *APOE* coding transcript. We propose that this transcript may be regulated by AD SNPs in a disease-state manner or could itself be driven by AD pathology. Long-read sequencing may be helpful to elucidate the full spectrum of the *APOE* transcripts in human brain tissue and human iPSC-derived brain organoids.

Despite the wealth of evidence linking *APOE* SNPs to pathology implicated in AD, an understanding of the specific mechanism(s) by which genetic variation at this region alters risk remains incomplete. *APOE* acts in conjunction with other genetic and environmental factors to confer AD risk. DNA methylation and chromatin status are associated with genetic and environmental factors, and previous studies have identified associations with AD and neuropathological hallmarks of AD in large collections of human brain tissue samples^36,37^. However, DNA methylation at the *APOE* locus has not been well studied. We found *APOE* alleles associated with AD and associated simultaneously with methylation levels of cg24084606, which was also reported to be involved in the autism spectrum. However, there was a weak association in a South African Cohort^38^.

Our data suggests that EGR4 and VDR might play a role during *APOE* gene transcription. EGR4, a zinc-finger transcription factor, is downregulated in AD mouse models’ brain^39^. It plays an important role in the developmental upregulation of KCC2 gene expression that is essential for fast synaptic inhibition in adult neurons^40^. Vitamin D can upregulate VDR^41^ and purportedly protect against cognitive decline and dementia^42^. However, the binding of one TF alone is rarely enough to directly infer functional effects on the gene expression levels, typically under the combinatorial and dynamic control of multiple TFs. Therefore, TF data are often actively integrated with other functional genomic techniques to decipher the basic regulatory control of gene expression, such as by incorporating active chromatin regions, DNA methylations, and SNPs. Interestingly, the SNP we prioritized is located within ChIP-seq peaks, correlated with CpG methylation levels, influences *APOE* jxn 1.2.2 transcript expression, and has genetic effects on AD core features in CSF (Aβ42 and pTau).

Our study revealed new *APOE* gene regulatory mechanisms affecting common AD risk SNP that may interact with chromatin, TFs, and DNA methylation to be responsible for turning the *APOE* transcription on or off in a different set of cells, or at different times. Though we identified potential functional variant associated with AD in this study, we still do not know how this genetic control of gene expression confers AD risk and pathology. It is likely that the identified SNP affects the *APOE* jxn1.2.2 expression level no matter the APOE genotype, and the change of *APOE* jxn1.2.2 expression may play a pivotal role in neuropathogenesis. We plan to assess whether repression of jxn1.2.2 expression through CRISPR assays in human induced pluripotent stem cell lines derived from rs157580-A carriers modulate AD-relevant phenotypes. If validated, these cell lines could then serve as models to test molecules as potential therapeutic interventions for treating rs157580-A carriers by manipulating the gene expression of *APOE* jxn1.2.2. Finally, this work also highlights the importance of including different ancestries in research on AD, as shared functional elements can provide windows of opportunity to cure the disease in diverse populations.

## MATERIALS AND METHODS

### ROSMAP

#### WGS Data processing

Whole-genome sequencing (WGS) datasets collected by the ROSMAP consortium were obtained from AD Knowledge Portal. There were 43,012,378 genomic variants in the raw data. Genetic variants were filtered out with PLINK 1.9^43^ if they: (1) had a genotype missing rate > 10%; (2) had Minor Allele Frequencies (MAF) < 1%; and (3) deviated from Hardy–Weinberg Equilibrium (HWE, *p-value* < 1E−6). Finally, we retained 9,912,554 common single nucleotide polymorphisms (SNPs) (23% of the total genetic variants).

#### IBD and PCA

To detect genetically related samples and population stratification, we applied stricter Quality Control (QC) procedures before conducting the Identity-By-Decent (IBD) test and Principal Component Analysis (PCA). First, we merged the study data with HapMap3 data and kept only the overlapped SNPs. We then removed SNPs if they: (1) had a genotype missing rate > 1%; (2) had MAF < 5%; (3) deviated from HWE (p-value < 1E−3), and (4) were in Major Histocompatibility Complex (MHC) regions (chr6:25M-33.5M). Finally, we retained 995,871 variants for further analysis. Pruning was conducted twice using PLINK with option --indep-pairwise 200 100 0.2. IBD test was conducted using PLINK with option --genome. Subjects with PI-HAT>0.2 were identified as the related subjects, and one of the related subjects with a higher overall SNP missing rate of the pair was removed. PCA was conducted with EIGENSOFT 6.1.3^44^. Twenty PCs were kept. Outliers of the population were detected in a training-prediction approach. We classified HapMap3 samples into two groups: EUR (CEU, TSI) and others. Next, we used 20 PCs of HapMap samples to fit a general linear model with glmnet, and then we used an estimated model to predict the probability of ancestry (ancestry score) for the studying sample. Subjects with ancestry scores lower than 0.8 were removed from study samples.

#### Bulk brain RNA-Seq data processing

Three brain regions of postmortem data were included in this study. The protocol of sample procurement has been described previously^13,45^. QC of the sequence data, including checks for over-abundance of adaptors and over-represented sequence, was performed using FastQC. Low-quality reads (5% of the total) were filtered out using the Trimmomatic^46^, which is a fast, multithreaded command line tool to trim and crop FASTQ data and remove adapters^46^. After trimming adapter sequences, reads passing initial QC were aligned to the human reference genome using HISAT2^47^ Gene lengths were calculated using GENCODE v41 annotations^48^. We converted gene counts to RPKM values using the total number of aligned reads across the 22 autosomal chromosomes. We converted junction counts to normalized values using the total number of aligned reads across the autosomal chromosomes, which can be interpreted as the number of reads supporting the junction in an average library size^15^.

#### *cis*-acting eQTL analysis

*cis*-eQTL association was examined separately by feature type (gene and junction) using TensorQTL package^22^, taking log2-transformed expression levels of each measurement (RPKM and RP10M) as the income. Features with low expression (average counts < 0.4 in gene and < 0.1 in junction) were excluded before eQTL analysis. To control for potential confounding factors, we adjust for ancestry (first 5 PCs) from the genotype data, diagnosis, sex, and the first K PCs of the normalized expression features, where K was calculated separately by feature type using the sva Bioconductor package^49^ (DLPFC: gene - 23 PCs, junction - 27 PCs). False discovery rate (FDR) was assessed across all *cis*-eQTL tests within each chromosome using R package qvalue^50^. We considered all variant–gene pairs (expression features to genes, eGene) and variant–junction pairs (eJunction) when the distance between features and SNP is <1MB.

#### Conditional analysis on APOE2,3,4 determining SNPs

We evaluated the effects of *APOE* loci on associations of candidate SNPs with the expression of APOE and the corresponding junctions. Since we don’t have data for APOE4 diplotypes in the LIBD sample, we used two *APOE* SNPs (rs7412 and rs429358) as covariates in the model to investigate the conditional effect of APOE2,3,4 alleles.

*fit0 = glm(expression ∼ Dx + Age + Sex + RIN + rRNA rate + totalAssignedGene + 5 SNP PCs + K feature PCs + rs7412 + rs429358, data=candi) Residual = resid(fit0)*

*fit1 = glm(residual ∼ SNP, data=candi*)

If the p-values of candidate SNPs in fit1 keep significance as original models without two *APOE* SNPs, we concluded that the effect of candidate SNPs is independent with two *APOE* SNPs.

#### Epistasis of candidate SNPs and APOE4 on expression

We used a general linear model and likelihood ratio test to evaluate the epistasis between the APOE4 haplotype and our candidate SNPs.

*Fit0 = glm(expression ∼ Dx + Age + Sex + RIN + rRNA rate + totalAssignedGene + 5 SNP PCs*

*+ K feature PCs, data=candi) Residual = resid(fit0)*

*fit1 = glm(residual ∼ SNP + APOE4, data=candi) fit2 = glm(residual ∼ SNP * APOE4, data=candi) lrtest(fit2, fit1)*

Results of the likelihood ratio test showed if there is an interaction effect of explanatory variables on response variables.

#### Differential Expression Analysis

Since we focused on only APOE and the related junctions, we used a general linear model to investigate the differential expression in diagnosis groups. We first fit a general linear model using Sex, Age, RIN, rRNA-Rate, the total number of assigned genes, 5 SNP PCs, and K number gene PCs used in eQTL analysis to keep consistency. We took the residual as the adjusted expression levels for further examination. Using the adjusted expressions, we conducted an ANOVA test using Anova in R to evaluate the difference between diagnosis groups. We also used the adjusted expressions for the related plots.

- *H*_0_: *μ*control=*μ*case
- *H_1_: mean is different*

*Fit0 = glm(expression ∼ Age + Sex + RIN + rRNA rate + totalAssignedGene + 5 SNP PCs + K feature PCs*

*Fit1 = glm(residual ∼ Dx) Result = Anova(fit)*

#### DNA Methylation Data processing

Methylation data was created on prefrontal cortex samples collected from 743 individuals using the Illumina HumanMethylation450 BeadChip by the ROSMAP consortium. After matching to QCed genotype data, we got 667 samples. QC and normalization were conducted using minfi R package^51^. Failed positions were identified with detectionP function in minfi by examining both the methylated and unmethylated channel reporting background signal levels. P-value for every genomic position in every sample was estimated. Small p-values indicate a good position. We excluded samples with averaged p-values > 0.05 across all probes, and also removed probes with averaged p-values > 0.05 across all samples. Normalization was conducted with function preprocessQuantile. We excluded probes on sex chromosomes to focus on mQTLs analysis on autosome chromosomes. We also removed probes that have the same locations as SNPs.

#### mQTL analysis

*cis*-mQTL association was examined for CpG using TensorQTL package^22^. To control for potential confounding factors, we adjusted for ancestry using the first five PCs from the genotype data, sex, and the first 2 Negative control PCs that were calculated with R Bioconductor package sva^21^ using QCed methylation data. FDR was assessed in R package qvalue^50^ across all QTL tests within each chromosome. We considered all variant–CpG pairs when the distance between CpG and SNP is <1MB.

#### ChIP-Seq data processing

Trim Galore was used to check the quality of the FASTQ files and run trimming. Bowtie 2 was used to align FASTQ files while the output was converted to the SAM file format. Samtools view was used to convert SAM files to BAM format. Bedtools intersect function was used to remove chrM, chrUN, pcr dup done with parameters, where blacklist is a list of unwanted sequences from the human reference genome. This output was then sorted using Samtools sort and potential PCR duplicates were removed using Samtools rmdup. To create bigWig file formats, deepTools bamCoverage was used. To obtain DNA binding motifs, we used Motif Scan and Enrichment Analysis (MoSEA) to scan for motifs. MoSEA can search for motifs against specified position weight matrices (PWMs). We used the HOmo sapiens COmprehensive MOdel COllection (HOCOMOCO) v11 mononucleotide in MEME format as the PWMs. MoSEA also incorporates MEME Suite’s Find Individual Motif Occurrences (FIMO)^52^ tool to scan for sets of sequences for individual matches to all motifs in HOCOMOCO v11^53^.

### LIBD

#### Genotype Data processing

SNP genotyping with HumanHap650Y_V3, Human 1M-Duo_V3, and Omni5 BeadChips (Illumina, San Diego, CA) was conducted with DNA extracted from brain cerebellar tissue. Genotype imputation was performed on TOPMed server with the imputation reference from the Human Reference Forum (https://topmedimpute.readthedocs.io/en/latest/). We retained common SNPs (MAF > 5%) that were present in the majority of samples (missingness < 5%) that were in HWE (p-value > 1 × 10^−6^) using the PLINK 1.9^43^. 9,984,191 SNPs were retained after QC.

#### Bulk brain RNA-Seq data processing

DLPFC RNA-Seq data from postmortem brain samples were included in this study. Details of tissue acquisition, handling, processing, dissection, clinical characterization, diagnoses, neuropathological examinations, RNA extraction, and quality control measures were described previously^54^. RNA extraction, sequencing, and RNA data processing were also described previously^15^. In our analysis, gene lengths were calculated using GENCODE v41 annotations^48^. We normalized gene counts and junction counts using the same approach as we did for ROSMAP data.

#### Cis-eQTLs analysis

*cis*-eQTL association was examined using TensorQTL package^22^, taking log2-transformed expression levels of each measurement (RPKM and RP10M) as the income. Features with low expression (average counts < 0.4 in gene and < 0.1 in junction) were excluded before eQTL analysis. To control for potential confounding factors, we adjust for ancestry (first five PCs from the genotype data), diagnosis, sex, and the first K PCs of the normalized expression features, where K was calculated separately by feature type using the sva Bioconductor package^49^ (AA: gene - 16, junction - 13).

## Data Availability

All data produced in the present work are contained in the manuscript

## ACKNOWLEDGEMENTS

The ROSMAP was supported by the National Institute on Aging (NIA) RF1AG57473, P30AG010161, R01AG015819, R01AG017917, U01AG46152, U01AG61356, RF1AG059082, P30AG072975, and R01AG036042. The authors would like to acknowledge NIA P30AG066546, U01 AG058589, R01 AG061872, U01 AG052409, R01 AG059421. The authors also acknowledge Bill and Rebecca Reed Endowment for Precision and Palliative Medicine. We acknowledge the Texas Advanced Computing Center (TACC) and Genomics, Epigenomics, Network, Imaging, and Education (GENIE) for providing high-performance computing (HPC) resources.

---

**Supplementary Figure S1.**
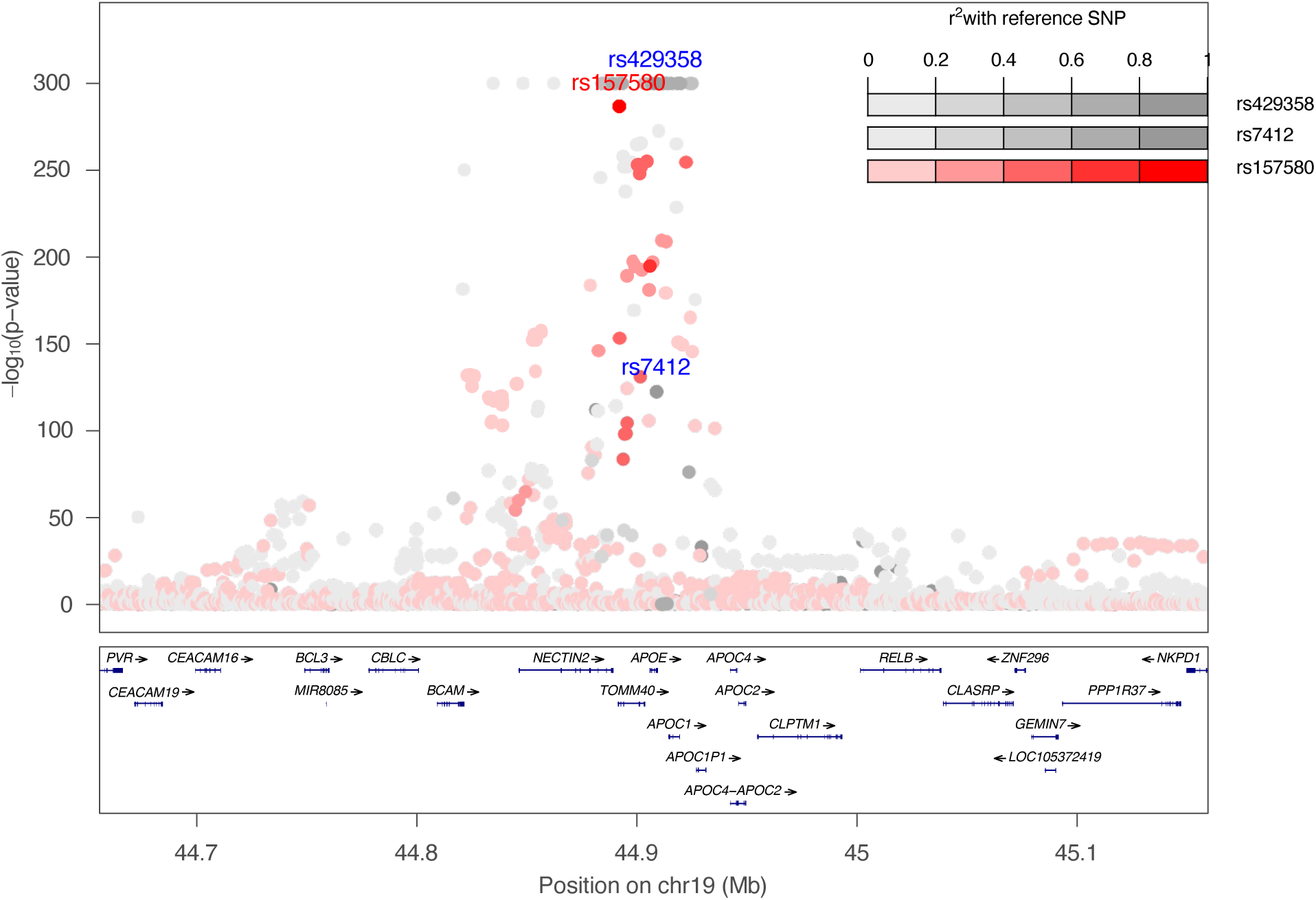
GWAS summary statistics at the APOE locus (hg38, chr19:44,655,791−45,159,393). Color is coded for linkage disequilibrium of predicted functional SNP rs157580 and SNPs consisting of APOE2,3,4 genotypes (rs429358 and rs7412). Minimum p-value = 1e-300.

**Supplementary Figure S2.**
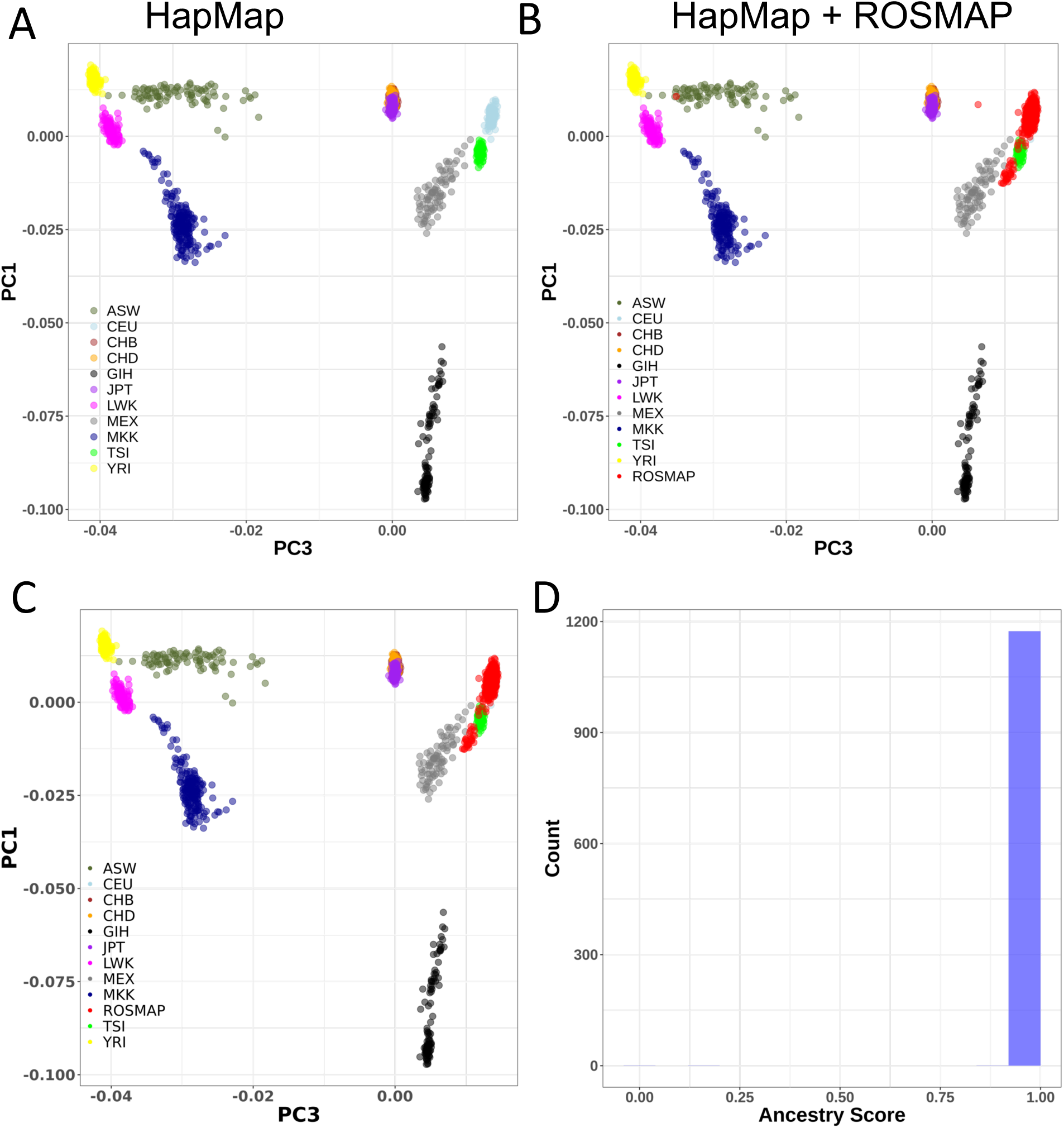
Principal component analysis (PCA) for (A) HapMap populations (reference), (B) ROSMAP European ancestry. (C) PC plots after removing outliers. (D) The ancestry score shows our populations are homogenous after removing outliers.

**Supplementary Figure S3.**
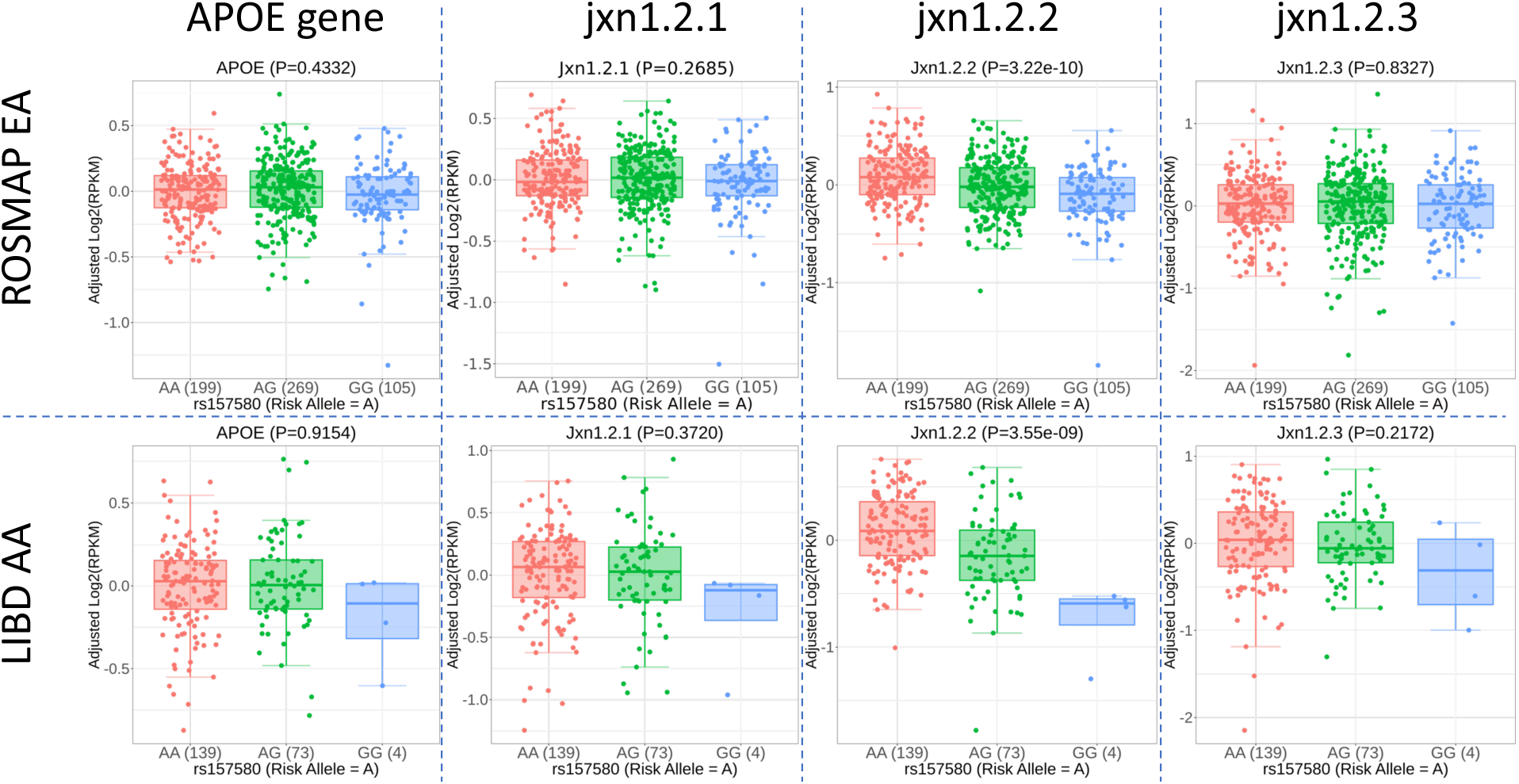
Conditional analysis of APOE expression features with rs157580. EA, European ancestry; AA, African American

**Supplementary Figure S4.**
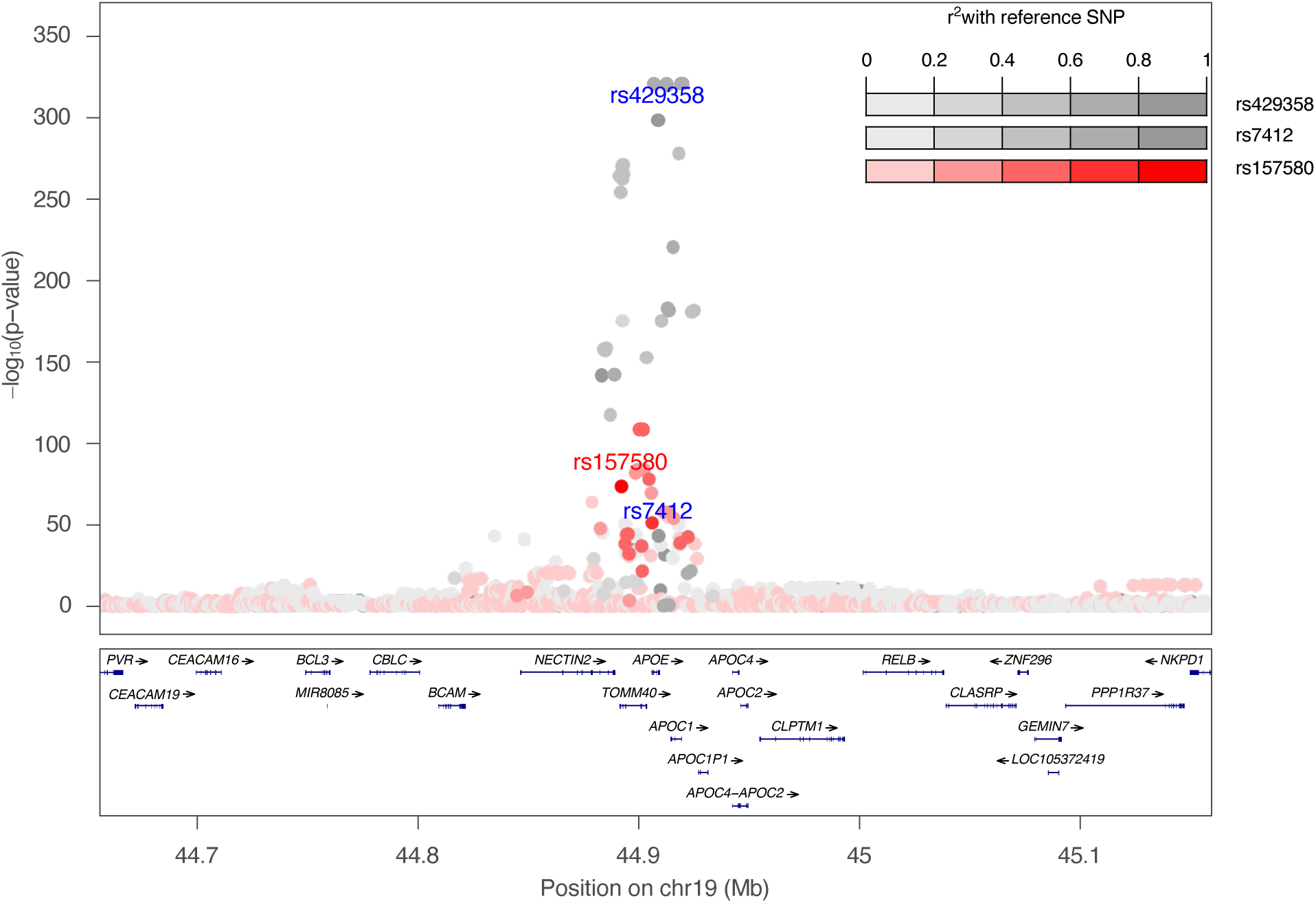
A-beta protein GWAS summary statistics at the APOE locus (hg38, chr19:44,655,791−45,159,393). Color is coded for linkage disequilibrium of predicted functional SNP rs157580 and SNPs consisting of APOE2,3,4 genotypes (rs429358 and rs7412). Minimum p-value = 1e-321.

**Supplementary Table S1.** The association between rs157580 and APOE transcripts is not influenced by APOE2 and APOE4 genotypes.

| **SNP** | **Transcript** | **p-value** | **FDR** | **p-value**<br>**(SNP X APOE4)** | **p-value**<br>**(SNP X APOE2)** |
| --- | --- | --- | --- | --- | --- |
| rs157580 | Jxn1.2.1 | 0.2685 | 0.9937 | 0.9195 | 0.1894 |
| rs157580 | Jxn1.2.2 | 1.71E-13 | 5.28E-10 | 0.1569 | 0.6391 |
| rs157580 | Jxn1.2.3 | 0.4621 | 0.9360 | 0.5110 | 0.3401 |
| rs429358 | Jxn1.2.1 | 0.5241 | 0.9937 | - | - |
| rs7412 | Jxn1.2.1 | 0.5720 | 0.9937 | - | - |
| rs429358 | Jxn1.2.2 | 1.99E-04 | 0.0228 | - | - |
| rs7412 | Jxn1.2.2 | 0.0827 | 0.6425 | - | - |
| rs429358 | Jxn1.2.3 | 0.1948 | 0.8044 | - | - |
| rs7412 | Jxn1.2.3 | 0.0927 | 0.7609 | - | - |

**NOTE:**
rs429358 and rs7412 are APOE2,3,4-determining SNPs.
Data is from ROSMAP DLPFC brain region.

